# The architecture of co-morbidity networks of physical and mental health conditions in military veterans

**DOI:** 10.1101/2020.06.10.20067116

**Authors:** Aaron F Alexander-Bloch, Armin Raznahan, Russell T Shinohara, Samuel R Mathias, Harini Bathulapalli, Ish P Bhalla, Joseph Goulet, Theodore Satterthwaite, Danielle S. Bassett, David C Glahn, Cynthia A. Brandt

## Abstract

Co-morbidity between medical and psychiatric conditions is commonly considered between individual pairs of conditions. However, an important alternative is to consider all conditions as part of a co-morbidity network, which encompasses all interactions between patients and a healthcare system. Analysis of co-morbidity networks could detect and quantify general tendencies not observed by smaller-scale studies. Here, we investigate the co-morbidity network derived from longitudinal healthcare records from approximately 1-million U.S. military veterans, a population disproportionately impacted by psychiatric morbidity and psychological trauma. Network analyses revealed marked and heterogenous patterns of co-morbidity, including a multi-scale community structure composed of groups of commonly co-morbid conditions. Psychiatric conditions including posttraumatic stress disorder were strong predictors of future medical morbidity. Neurological conditions and conditions associated with chronic pain were particularly highly co-morbid with psychiatric conditions. Across conditions, the degree of co-morbidity was positively associated with mortality. Co-morbidity was modified by biological sex and could be used to predict future diagnostic status, with out-of-sample prediction accuracy of 90-92%. Understanding complex patterns of disease co-morbidity has the potential to lead to improved designs of systems of care and the development of targeted interventions that consider the broader context of mental and physical health.

## Introduction

More than 25% of U.S. citizens have multiple chronic medical conditions, and these individuals drive the majority of healthcare costs^1^. Analogously, psychiatric conditions are not epidemiologically independent, but co-occur throughout much of the population. This phenomenon clearly impacts clinical care, treatment efficacy, likelihood of polypharmacy, and risk of redundant testing and intervention. The complexity that accompanies comorbid conditions is only compounded when considering the relationships between psychiatric multimorbidity and medical conditions^3^. For example, posttraumatic stress disorder has been linked to multiple chronic medical conditions in US military veterans^4-6^. The complexity of the set of co-morbidity relationships suggests its suitability for analysis as a network, as the growing field of network science has developed to study precisely this kind of multivariate complexity^7^, in order to identify organizing principles that could explain the co-occurrence of conditions within individuals over time.

In addition to its public health implications, the extent of psychiatric co-morbidity is often considered to be a major limitation of existing diagnostic classification systems, driving the need for the scientific development of more biologically-informed classifications^8,9^. Psychiatric comorbidity is partly due to (sometimes trivial) overlap in diagnostic criteria^10,11^ and may also be partly due to a single psychiatric vulnerability “p factor” that underscores all psychopathology^12^. But multiple studies also show that co-morbidity relationships recapitulate specific, shared genetic risk between clinical conditions^13-15^ and cellular protein-protein or metabolic pathways linked to disease pathogenesis^16-18^. Electronic health records (EHRs) containing diagnostic information can thus be leveraged to provide preliminary data about co-morbidity relationships worthy of further investigation.

The U.S. veteran’s health administration (VA) has one the largest EHRs for an analysis of comorbidity relationships. Moreover, because of their unique exposure to psychological trauma especially in the post-9-11 era, veterans represent a singular resource for the investigation of psychiatric co-morbidity networks, particularly as they relate to post-traumatic stress disorder (PTSD). Previous work has suggested significant co-morbidity between PTSD and other conditions including alcohol use disorder, depression, cardiovascular disease, and chronic pain^4-6^. A significant national investment in an extensive VA EHR^19^ allows patterns of medical and psychiatric co-morbidity to be systematically evaluated in this population, to move beyond studies of individual conditions.

Here, we use a network science approach to map the relationships between 95 psychiatric and medical conditions, with health records from approximately 1 million U.S. military veterans. In multiple ways, the present study extends existing work on the analysis of co-morbidity networks. First, we specifically investigate the relationship between psychiatric and medical conditions, showing broad co-morbidity that is particularly strong with neurological conditions and conditions associated with chronic pain. Second, we explicitly model the temporal aspect of comorbidity, showing that psychiatric conditions including PTSD function as predictors of future medical morbidity. Third, we demonstrate the clinical utility of leveraging network-level comorbidity information into a clinical vulnerability score, which could be used to flag individuals at risk for the development of schizophrenia and suicidal/self-harm behaviors. Finally, we replicate a prior result showing an association between the degree of co-morbidity and mortality across conditions. Together, results demonstrate that marked and heterogenous co-morbidity between physical and mental health has the potential to inform the design of systems of care, clinical practice, and biological research in the future.

## Materials and Methods

VA administrative and clinical data were derived from the VA National Corporate Data Warehouse, which provide coded diagnostic data associated with all VA inpatient and outpatient encounters. The study sample consisted of U.S. Veterans who served in Operation Enduring Freedom, Operation Iraqi Freedom, or Operation New Dawn (OEF/OIF/OND). Inclusion criteria were enrollment in VA healthcare after October 1, 2001 and having one or more VA primary care visits prior to the end of the study period on October 1, 2017. We excluded 310 individuals without a recorded birthdate, and an additional 15 with unknown sex (140 of the individuals without a recorded birthday also had unknown sex), yielding a final sample of 977,183 individuals. The study was approved by the Human Investigation Committees at VA Connecticut Healthcare System—West Haven.

### Medical and Psychiatric Conditions of Interest

The principal conditions were derived from a set of mappings of ICD9/10 codes to clinically meaningful conditions, created by the U.S. Agency for Healthcare Research and Quality (AHRQ) and previously used in many epidemiological and health service studies^20^ (https://www.hcup-us.ahrq.gov/toolssoftware/ccs/ccs.jsp#pubs). AHRQ conditions comprise four levels from broadest (level 1) to most specific (level 4). For example, “hypertensive heart and/or renal disease” (level 4) is contained within “hypertension with complications and secondary hypertension” (level 3), “hypertension” (level 2), and “diseases of the circulatory system” (level 1). To balance interpretability and specificity, we used the set of level 2 conditions for this analysis, providing a broad sampling across the diagnostic space.

In addition, the AHRQ conditions were supplemented with 9 mappings to psychiatric conditions provided by the VA’s Northeast Program Evaluation Center (NEPEC; https://www.ptsd.va.gov/about/divisions/evaluation/). All NEPEC conditions as well as the following AHRQ conditions were considered to be psychiatric: dementia, developmental disorder, early childhood disorders, personality disorder, suicide/self-injury, impulse control disorder, adjustment disorder, attention deficit disorders. AHRQ conditions that were redundant with NEPEC conditions were excluded, including: alcohol-related disorders, mood disorders, anxiety disorders, psychotic disorders, substance use disorders.

We further curated the AHRQ data in several additional steps. First, we removed the following AHRQ categories, which reflected healthcare utilization rather than conditions: care in pregnancy, contraceptive management, immunization, maintenance chemotherapy, and screening for mental health disorders. In addition, categories that were deemed *a priori* to be too broad for interpretability were excluded: “complications”, “miscellaneous mental health disorders”, “ill-defined symptoms”, “normal pregnancy”, and “complications related to pregnancy”. Finally, we combined 14 sub-categories of cancer/neoplasms into a single category, 5 sub-categories of congenital anomalies into a single category, diabetes with and without complications, and male/female genital disorders.

As previously described^21^, we counted a veteran as having any condition that was coded at least once for an inpatient stay or at least twice for an outpatient visit. This criterion was used because outpatient codes are assigned by health care providers and are expected to be less complete than inpatient codes, which are assigned by professional coders^21^. This methodology has been found to improve accuracy in the identification of psychiatric disorders in administrative data^22^ and in the identification of HIV in Medicaid data^23^. We also excluded disorders with prevalence lower than 0.1%, resulting in a total of 95 medical or psychiatric conditions.

### Statistical Modelling

For each pair of conditions, dx_i_ and dx_j_, we sought to determine whether the odds of diagnosis with disease dx_i_ were affected by diagnosis with dx_j_ (and vice versa). The basic statistical approach was the logistic regression model, which is used to model dichotomous outcome variables, implemented in the R *stats* library^24^. The outcome variable was diagnosis with a single condition of interest, dx_i_. The log odds of having the condition was modeled as a linear combination of the predictor variables. After the model is fit to the data, each coefficient, *β*, corresponds to the contribution of a predictor variable to the model, where the log odds of having the outcome condition increase by a factor of *β*. The following predictor variables were included in the model: diagnosis with dxj; multimorbidity, quantified as the total number of medical (non-psychiatric) conditions not including dx_i_ or dxj; P-factor, quantified as the total number of psychiatric conditions not including dx_i_ or dxj; linear, quadratic and cubic effects of age; sex; reported race, with categories including Caucasian, African American, Hispanic, and Other/Unknown according to VA records; and the number of years since final deployment. The number of years since final deployment is a proxy for years of opportunity to use VA services, which was included as a co-variate in the model because it could increase the likelihood of diagnosis with multiple conditions. For each *β*, P-values were calculated using Wald tests of the null hypothesis that *β* = 0^24^.

The inclusion of multimorbidity allowed us to disentangle co-morbidity from a general effect whereby sick people, who naturally utilize healthcare often, have more diseases and diagnostic encounters. Likewise, the P-factor accounts for a general effect where people with psychiatric vulnerability might have more psychiatric diagnoses. The null hypothesis with respect to dx_j_ is that having dx_j_ does not increase the probability of having dx_i_, above and beyond the probability predicted by the individual’s number of other medical or psychiatric conditions.

Critically, the above model was also modified to incorporate directionality in time, in order to focus on whether prior diagnosis with dx_j_ increases the odds of subsequent diagnosis with dx_i_. To incorporate this constraint, for the purposes of modelling the outcome variable dx_i_, dx_j_=1 if and only if the diagnosis of dx_j_ occurred prior to or on the same date as the diagnosis of dx_i_. The multimorbidity and P-factor scores were also adjusted, incorporating only the number of diagnoses that existed at the time of diagnosis with condition dx_i_. Note that with this methodological approach, the effect of dx_j_ on dx_i_ is not expected to be the same as the effect of dx_i_ on dx_j_.

### Network Analysis

In order to analyze patterns of co-morbidity across all conditions, we modelled the set of comorbidity relationships as a network rather than considering each pair of conditions separately. In general, a network is a mathematical model of elements (‘nodes’) and their interactions (‘edges’). Here, nodes were disease conditions, and edges were significant co-morbidity relationships between pairs of disease conditions estimated from the logistic regression models described in the previous section. Edges were included in the adjacency matrix, **A**, if there was a positive effect that was statistically significant at P<0.05 after FDR-correction for multiple comparisons. Specifically, the element *A_ij_* was the log odds increase of condition *i* given condition *j*.

To quantify the overall co-morbidity of a condition, we use the network property of strength. The strength of node *i, s_i_*, is the sum of the weight of all of its edges. To differentiate between 1) condition *i*’s impact on the odds of diagnosis with other conditions and 2) other conditions’ impact on the odds of diagnosis with condition *i*, strength was further divided into in-strength and out-strength. In-strength, 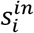, was the sum of edges directed towards node *i;* out strength, 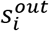, was the sum of edges directed away from node *i*. These two measures calculated for single nodes quantify the centrality of the node in terms of connectivity from or to other nodes^25^. A high value of 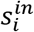 indicates that diagnoses of many other conditions are strongly predictive of future diagnosis with condition dx_j_, while high value of 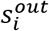 indicates that diagnosis with condition dx_j_. is strongly predictive of future diagnoses of many other conditions.

If similar conditions tend to have disproportionate co-morbidity with one another - greater than their co-morbidity with other nodes in the network - then these conditions may form a community within the overall network, which can be investigated as a semi-distinct network in its own right. To perform community detection, we employed the generalized Louvain algorithm implemented in MATLAB (http://netwiki.amath.unc.edu/GenLouvain) to maximize a modularity quality index defined separately for the psychiatric and medical co-morbidity networks^26^. The modularity quality function that we maximized to obtain a partition of nodes into communities is particular to directed networks and defined as follows^27^:

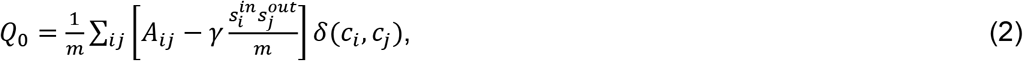

where *m* is the total weight of all edges in the network, and the delta function *δ*(*c_i_, c_j_*) = 1 if node *i* and node *j* are in the same community, and *δ*(*c_i_, c_j_*) = 0 otherwise. The resolution parameter, *γ*, biases the partition solution towards either a larger or smaller number of communities^28^. We used a *γ* range from 0 to 4 in 0.01 intervals, with 20 iterations of the algorithm at each resolution, in order to sample across possible resolutions. The stochastic maximization algorithm is repeated multiple times because the modularity function *Q* can exhibit multiple, distinct high-scoring solutions^29^.

It is important to sample across resolutions because it has been shown that even clearly-defined modular communities can be missed for a single value of *γ*^30^. Moreover, community structure is often present at multiple resolutions^31^. To identify representative community partitions, firstly, all partitions at a given resolution were matched by iteratively solving the linear assignment problem in order to generate a consensus partition at that resolution^32,33^. Representative community partitions were then chosen based on 1) stability of the partition across a relatively wide range of *γ* values in terms of the number of modules and ^2^) representativeness in terms of a relatively high normalized mutual information (NMI) with partitions at other resolutions^34,35^.

To quantify the co-morbidity between modules, we used the measure of normalized intermodule co-morbidity *(NIC). NIC* is defined as the sum of the inter-module edge weights from module A to module B, divided by the product of the number of nodes in each module.

### Pain, mortality and sex effects

We sought to specifically test the hypothesis that pain disorders have a particularly strong association with psychiatric conditions, either because physical pain causes psychological pain or because they share underlying mechanisms. In order to estimate the association with pain for the set of conditions used in this analysis, we used a recently-validated approach to map ICD9/10 codes to 15 common pain conditions^36^. As identified, the pain conditions were quite broad (e.g. limb pain, back pain, neck pain), many with definitional overlap with at least one of the AHRQ conditions. Therefore, rather than include the pain conditions in the comorbidity analysis, we identified a pain score for each AHRQ condition as the increase in odds of having that condition per number of pain conditions in separate logistic regression models that included the number of pain conditions rather than dx_j_ as a predictor variable. The pain scores were in turn used to assess whether the association with pain were distinct across the different network communities. To quantify the mortality of each condition, we used the increase in odds of each condition with the outcome variable of death during the study period, which occurred for n=17,317 veterans.

Given the increased clinical focus on women’s psychiatric health and the importance of sex as a biological variable of interest^37,38^, we sought to determine whether sex augments co-morbidity beyond sex-related differences in disease prevalence. Formally, we added an interaction effect, *dxj*sex*, to the logistic regression model as an additional predictor variable Statistical significance was determined using the Wald test with FDR correction for multiple comparisons.

### Predictive Potential

To assess the potential clinical utility of a network co-morbidity approach, we performed out-of-sample prediction of two psychiatric conditions of interest: schizophrenia and suicidal/self-injurious behaviors. The two conditions were chosen given the high clinical importance of their early prediction^39,40^. We randomly divided the data into a training set (80% of data) and a test set (20% of data). Using the training set, we performed lasso logistic regression using a model containing all diagnostic conditions implemented in the R package *glmnet*^41,42^. In all subjects, we excluded conditions that were diagnosed subsequent to the diagnosis of interest. For example, in the case of schizophrenia, lipid disorders diagnosed subsequent to the diagnosis of schizophrenia were not permitted to influence the model. The shrinkage parameter, *λ*, was chosen by 10-fold cross validation within the training set only. As the output of the model is probabilistic, ranging from 0 to 1, we empirically chose a probability cutoff for the purposes of prediction by maximizing Youden’s index^43^, *J* = Sensitivity + Specificity - 1, in the training data. Diagnostic status was then predicted for the test set using the model fitted to the training set data and compared to the actual diagnostic status to yield information about accuracy, specificity, and sensitivity. Receiver operating characteristic (ROC) curves for the models were compared to models derived solely from demographic information about subjects. Note that although the predictive models we describe do not make direct use of the co-morbidity network derived previously, it is the co-morbidity between all other conditions and the condition of interest that yields the predictive potential of this analysis.

## Results

The sample included a diverse cross-section of veterans, and co-morbidity was the rule rather than the exception. The mean age was 40.8 years (S.D. =9.62yrs), including 121,017 veterans whose reported sex was female and 856,166 veterans whose reported sex was male. Reported race comprised 158,431 African American; 109,121 Hispanic; 632,119 non-Hispanic Caucasian; and 77,512 veterans identified as another race. The mean date of first diagnosis was August 7, 2010 (S.D. = 2.6 years). The mean number of co-morbidities was 7.3 (S.D. = 4.1), and 77% of the veterans had been diagnosed with at least 2 of the conditions of interest. For veterans with at least 2 diagnoses at different visits, the mean difference between the times of first and last diagnoses was 2.3 years (S.D. = 2.2 years). The most frequently diagnosed psychiatric conditions were PTSD (36% of the sample), mild depression (symptoms that did not meet criteria for major depressive disorder, 28%), and unspecified anxiety (20%). Among the most frequent non-psychiatric diagnoses were nontraumatic joint disease (44%), back pain or spondylosis (42%), disorders of lipid metabolism (25%), hypertension (18%), and headache (18%). See Table 1.

**Table 1:** Cohort demographic information.

|  | **N** | **Mean** | **Standard Deviation** |
| --- | --- | --- | --- |
| **Age** |  | 40.8 yrs | 9.62 yrs |
| <35 yrs | 342,688 |  |  |
| 35-44 yrs | 310,350 |  |  |
| >44 yrs | 325,145 |  |  |
| **Reported Race** |  |  |  |
| African American | 158,431 |  |  |
| Hispanic | 109,121 |  |  |
| Caucasian | 632,119 |  |  |
| Other | 77,512 |  |  |
| **Reported Sex** |  |  |  |
| Female | 121,017 |  |  |
| Male | 856,166 |  |  |
| **Years in VA** |  | 7.5 yrs | 3.0 yrs |
| **Education** |  |  |  |
| <Highschool | 12,448 |  |  |
| Highschool | 219,948 |  |  |
| >Highschool | 744,787 |  |  |
| **Marital Status** |  |  |  |
| Married | 442,218 |  |  |
| Not married | 485,144 |  |  |
| Unknown | 420 |  |  |
| **Rank** |  |  |  |
| Enlisted | 893,174 |  |  |
| Commissioned Officer | 74,092 |  |  |
| Warrant Officer | 9,917 |  |  |
| **Earliest Diagnosis** |  | Aug 7, 2010 | 2.6 yrs |
| **Time between Diagnoses\*** |  | 1.2 yrs | 2.2 yrs |
| **Comorbidities** |  | 7.3 diagnoses | 4.1 diagnoses |
| 0 | 135,168 |  |  |
| 1 | 86,472 |  |  |
| 2 | 69,328 |  |  |
| 3 | 65,846 |  |  |
| 4 | 63,088 |  |  |
| >4 | 557,281 |  |  |

\*Difference between time of diagnosis with latest diagnosis and time of diagnosis with earliest diagnosis, for Veterans with at least 2 diagnoses at different times.

### Network architecture of psychiatric co-morbidity

Co-morbidity between psychiatric conditions is known to be high, possibly reflecting a psychiatric vulnerability factor that is generalized across conditions, or else more specific shared etiologies or vulnerabilities between certain conditions. As predicted, there was extremely high co-morbidity within the 17 psychiatric conditions. Out of 272 possible relationships tested, 100 were significant with an odds ratio greater than 1.5. Importantly, the statistical approach controlled for overall psychiatric morbidity, suggesting that pairwise co-morbidity was not due merely to a generalized psychiatric vulnerability factor.

The degree of psychiatric co-morbidity was not equal across disorders, being instead characterized by hierarchical community structure. Upon visual inspection, within the overall psychiatric co-morbidity network, there was evidence of communities with relatively denser connections, which were themselves composed of smaller, even more densely connected communities. (We here describe a community structure as “hierarchical”, a term with many meanings across disciplines, in reference to the existence of nested communities within communities.) Diagnostic plots (Supplemental Figure 1) show that the community structures at n=3 and n=6 modules were relatively stable and representative (see methods and Supplemental Figure 1), and these resolutions were investigated in greater detail. The 3-module partition included a developmental module (ADHD, developmental disorders and childhood disorders) and two relatively heterogenous modules: 1) a module that included schizophrenia, bipolar disorder, substance use disorders, and suicidality; 2) a module that included depression, anxiety, PTSD, and cognitive disorders. The larger modules contained smaller modules from the 6-module partition, including: a substance use module, with drug and alcohol use disorder; a depression/anxiety module with MDD, minor depression, GAD and other anxiety, as well as adjustment disorders; a psychosis module containing schizophrenia and bipolar disorder; and a module including suicidality, personality disorders, and impulse control disorders (see Figure 1 and Supplemental Figure 1).

**Figure 1:**
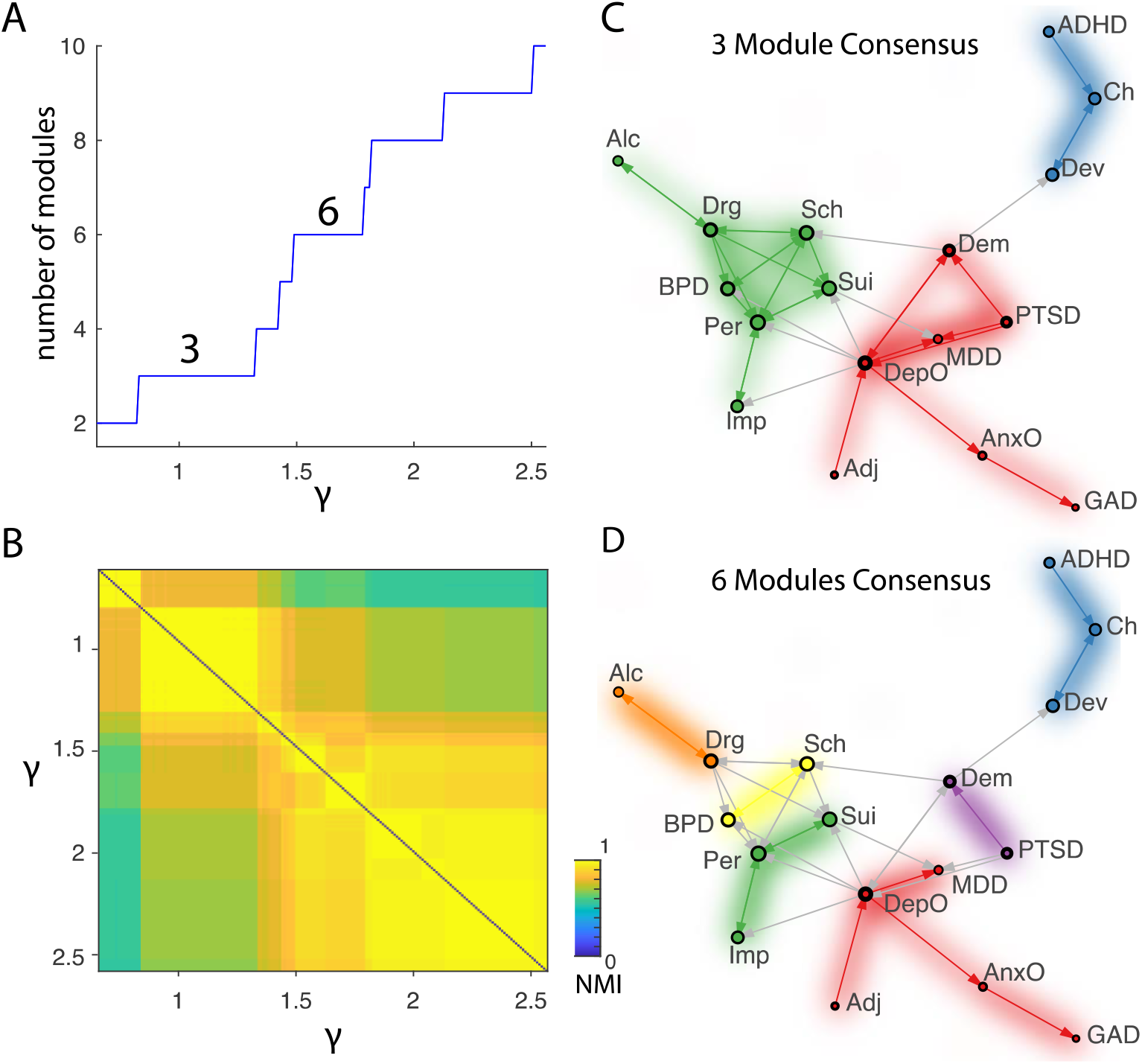
Illustration of the community structure of the psychiatric co-morbidity network. A) Diagnostic plots showing the number of modules at different resolution, quantified by the parameter *γ* from Equation (1) and B) the normalized mutual information (NMI) of the community structure at some given resolution and the community structure at another resolution. C-D) Networks represented with a selection of illustrative edges (the minimum spanning tree and the 10% highest-weight edges) using a force-directed representation with communities labelled by color (https://www.brainnetworkslab.com/coderesources). Arrows go towards the predicted condition. Inter-modular edges are colored gray. For the nodes, the area of the colored inner-circle is proportional to the in-strength, while the area of the outer black ring is proportional to the out-strength. Two relatively stable resolutions, with n=3 modules (C) and n=6 modules (D) are shown, using the consensus partition of all partitions at this resolution. See Table 2 for list of abbreviations.

**Table 2:** List of abbreviations of conditions.

| Abbr | Total n (x1000) | Module<sup>1</sup> | Condition (AHRQ full name) | Abbr | Total n (x1000) | Module<sup>1</sup> | Condition (AHRQ full name) |
| --- | --- | --- | --- | --- | --- | --- | --- |
| ADHD | 37 | PSYCH | Attention deficit hyperactivity disorder | Liv | 58 | HEM/CIRC | Liver disorder |
| Adj | 145 | PSYCH | Adjustment disorder | LRO | 104 | RESP | Lower respiratory disorder* |
| Alc | 81 | PSYCH | Alcohol use disorder | MDD | 186 | PSYCH | Major depressive disorder |
| Ane | 34 | HEM/CIRC | Anemia | Mou | 10 | RESP | Mouth disorder |
| AnxO | 192 | PSYCH | Anxiety* | Myc | 58 | RESP | Mycoses |
| Arl | 2 | ID | Infective arthritis | Neo | 95 | GI | Neoplasm or malignancy |
| Art | 69 | HEM/CIRC | Disorder of arterial system | NIn | 1 | NEURO | Central nervous system Infection |
| Ast | 45 | RESP | Asthma | NSC | 21 | NEURO | Hereditary and degenerative nervous system conditions* |
| Bac | 22 | ID | Bacterial Infection | NSO | 172 | NEURO | Nervous system disorders* |
| Back | 409 | MSK | Back pain or spondylosis | NTJ | 435 | MSK | Nontraumatic joint disorder |
| Blj | 72 | NEURO | Intracranial Injury | NuO | 224 | ENDO/METAB | Nutritional, endocrine or metabolic disorder* |
| Bil | 9 | HEM/CIRC | Biliary Tract Disorder | Nut | 73 | ENDO/METAB | Nutritional deficiency* |
| Bone | 36 | MSK | Bone disorder* | Ost | 2 | MSK | Osteoporosis |
| BPD | 32 | PSYCH | Bipolar Disorder | Pan | 4 | HEM/CIRC | Pancreatic disorder |
| Bur | 3 | ID | Burns | Par | 3 | NEURO | Paralysis |
| Ch | 2 | PSYCH | Childhood Psychiatric Disorder (including elimination and tic disorders) | Per | 23 | PSYCH | Personality disorder |
| Clj | 3 | MSK | Crushing/internal injury | PFr | 1 | MSK | Pathological fracture |
| Coag | 7 | HEM/CIRC | Coagulation Disorder | Ple | 4 | HEM/CIRC | Pleurisy, pneumothorax or pulmonary collapse |
| Coma | 2 | NEURO | Coma or brain damage | Poi | 6 | HEM/CIRC | Poisoning |
| Conn | 249 | MSK | Connective tissue disorder* | PTSD | 354 | PSYCH | Posttraumatic stress disorder |
| Cong | 22 | MSK | Congenital Anomaly | RF | 3 | HEM/CIRC | Respiratory Failure |
| COPD | 21 | RESP | COPD or bronchiectasis | RIn | 147 | RESP | Respiratory Infection |
| CVD | 8 | NEURO | Cerebrovascular Disorder | Sch | 8 | PSYCH | Schizophrenia or schizoaffective disorder |
| Def | 65 | MSK | Acquired Deformity | SIn | 43 | ID | Skin Infections |
| Dem | 33 | PSYCH | Dementia or other cognitive disorder | SInf | 33 | RESP | Inflammatory condition of skin |
| DepO | 272 | PSYCH | Minor Depression | SKO | 166 | RESP | Skin disorder* |
| Dev | 4 | PSYCH | Developmental disorder (including communication disorders and intellectual disability) | Spi | 1 | NEURO | Spinal cord injury |
| DM | 79 | ENDO/METAB | Diabetes mellitus | Spr | 88 | MSK | Sprain or strain |
| Drg | 84 | PSYCH | Substance use disorder (other than alcohol) | Sui | 37 | PSYCH | Suicide or self-injurious behavior |
| Ear | 279 | MSK | Ear Conditions | Sulj | 30 | MSK | Superficial injury |
| Ele | 21 | HEM/CIRC | Fluid or electrolyte disorder | Thy | 6 | ENDO/METAB | Thyroid disorder |
| End | 26 | ENDO/METAB | Endocrine Disorder* | TJ | 65 | MSK | Traumatic joint disorder |
| Ep | 14 | NEURO | Epilepsy or convulsions | UGI | 164 | GI | Upper gastrointestinal disorder |
| Eye | 255 | RESP | Eye Condition | Ulc | 2 | ID | Chronic skin ulcer |
| Fr | 49 | MSK | Fractures | Uri | 97 | HEM | Urinary system disorder |
| GAD | 44 | PSYCH | Generalized anxiety disorder | URO | 132 | RESP | Upper respiratory disorder* |
| Gas | 12 | GI | Noninfectious gastroenteritis | Vei | 48 | GI | Disorder of venous system |
| Gen | 140 | ENDO/METAB | Genital disorder | Vir | 61 | RESP | Viral infection |
| GIH | 30 | GI | Gastrointestinal hemorrhage | WBC | 14 | HEM/CIRC | Disorder of white blood cells |
| GIO | 103 | GI | Gastrointestinal disorder* | Wou | 19 | ID | Open wounds |
| Gou | 14 | ENDO/METAB | Gout or other crystal arthropathy |  |  |  |  |
| HA | 174 | NEURO | Headache |  |  |  |  |
| Hea | 122 | HEM/CIRC | Disorder of the heart |  |  |  |  |
| HeO | 4 | HEM/CIRC | Hematologic Condition* |  |  |  |  |
| Her | 31 | GI | Abdominal Hernia |  |  |  |  |
| HTN | 175 | ENDO/METAB | Hypertension |  |  |  |  |
| IIn | 5 | GI | Intestinal Infection |  |  |  |  |
| IjO | 67 | MSK | External Injuries* |  |  |  |  |
| Imm | 1 | RESP | Immunity disorder |  |  |  |  |
| Imp | 9 | PSYCH | Impulse control disorder |  |  |  |  |
| InO | 7 | RESP | Infection* |  |  |  |  |
| Jaw | 215 | RESP | Tooth or jaw disorder |  |  |  |  |
| LGI | 30 | GI | Lower gastrointestinal disorder |  |  |  |  |

Abbr, Abbreviation used for condition in figure labels; AHRQ, U.S. Agency for Healthcare Research and Quality; Psych, psychiatry module; GI, gastrointestinal module; MSK, musculoskeletal module; HEM/CIRC, a hematologic/circulatory system module; RESP, a respiratory module; ID, infectious disease module; NEURO, neurological module ; ENDO/METAB, endocrine/metabolic module <sup>1</sup>Community membership assigned by community detection algorithm (7-module solution; module name does not necessarily align with standard medical classification)

In addition to community membership, a major distinguishing factor between the psychiatric conditions was their relative in-strength and out-strength, which tracked their role as predictors of further psychiatric conditions in the future (out-strength) versus their own predictability based on prior psychiatric conditions (in-strength). Evidently, comorbid conditions are not always diagnosed simultaneously, and comparing the time of first diagnosis in patients with two conditions showed the paths that veterans tend to traverse through the psychiatric co-morbidity network. The relevance of the temporal dynamics is underscored by the fact that 38 of the 100 edges with odds ratios greater than 1.5 were not reciprocal, meaning that a predictive relationship from disease A to disease B did not necessarily imply a reciprocal relationship from disease B to disease A. Minor depression, PTSD, and dementia in particular were distinguished by their role as predictors, with their out-strength more than double their in-strength. In contrast, alcohol use disorder and ADHD were marked by their relative predictability from other conditions (see Figure 1 and Supplemental Figure 2).

### Community structure of medical co-morbidity

Co-morbidity between medical conditions was also high and, like that of psychiatric conditions, showed a strong hierarchical community structure (Figure 2, Supplemental Figure 3). A 7-module partition was relatively stable, representative, and mapped onto clinically relevant phenotypes including: a gastrointestinal module (GI), a musculoskeletal module (MSK), a hematologic/circulatory system module (HEM/CIRC), a respiratory module (RESP), and infectious disease module (ID), a neurological module (NEURO), and an endocrine/metabolic module (ENDO/METAB). These modules contained within them smaller modules from higher resolution partitions such as the 15-module partition also illustrated in Figure 2.

**Figure 2:**
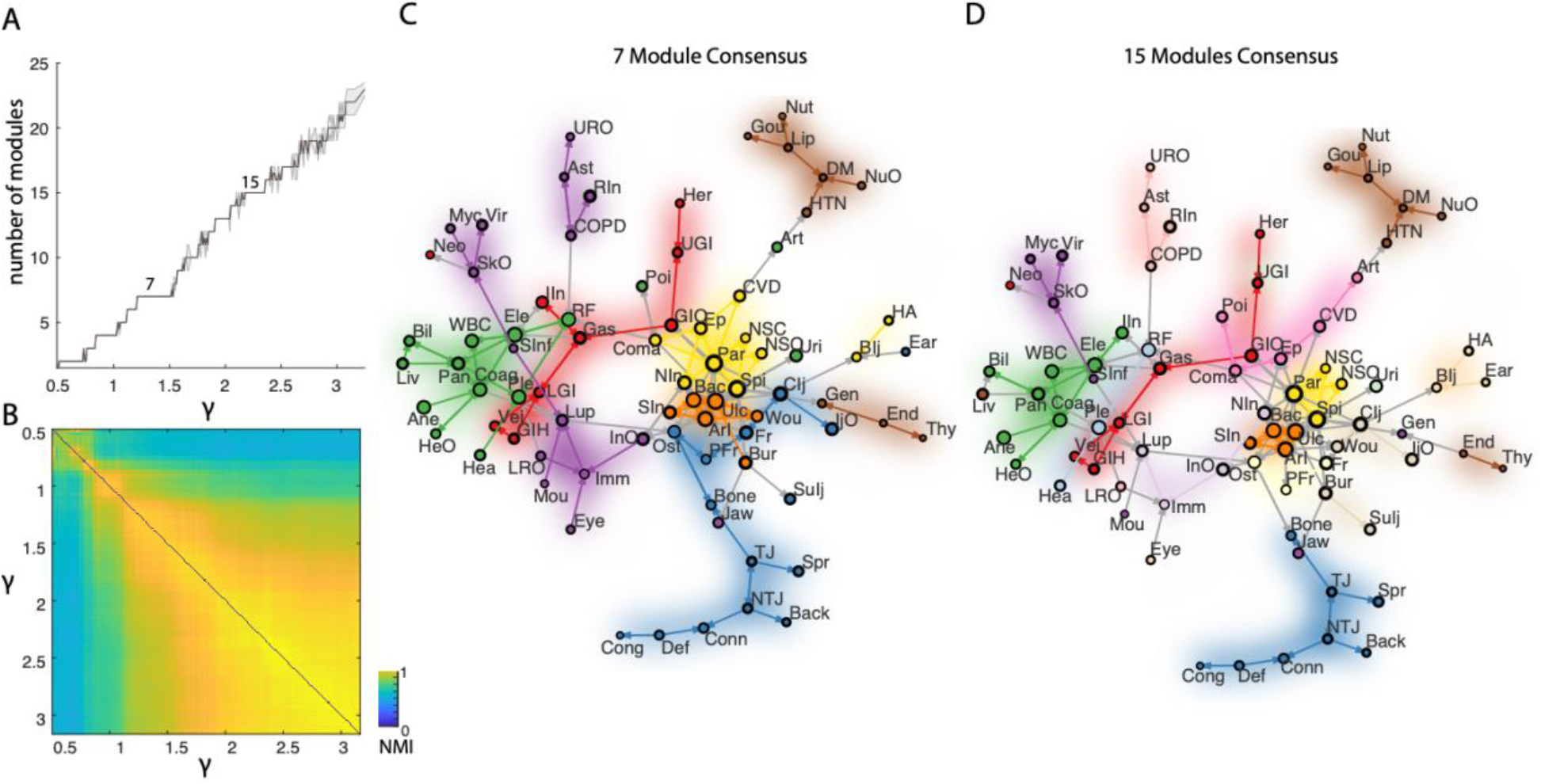
Illustration of the community structure of the medical co-morbidity network. A) Diagnostic plots showing the number of modules at different resolutions, quantified by the parameter y from Equation (1) and B) the normalized mutual information (NMI) with the community structure at other resolutions. C-D) Networks represented with a selection of illustrative edges (the minimum spanning tree and the 2% highest-weight edges) using a force-directed representation with communities labelled by color (https://www.brainnetworkslab.com/coderesources). Arrows go towards the predicted condition. Two relatively stable resolutions, with n=7 modules (C) and n=15 modules (D) are shown, using the consensus partition of all partitions at these resolutions. See Table 2 for list of abbreviations.

When considering the complete network of both psychiatric and medical conditions, we expected psychiatric and medical conditions to be mutually predictive, in terms of medical conditions increasing the odds of psychiatric conditions and vice versa. In fact, there was a clear bias in the direction of psychiatric conditions being indicators of future medical morbidity (Figure 3). Quantified by the normalized inter-module co-morbidity (*NIC*, see methods) and treating the psychiatric conditions as a single module for the purpose of comparison with the medical modules, the psychiatry module had a larger predictive effect on future diagnosis with medical conditions, compared to the reciprocal effect of medical conditions on the odds of developing a future psychiatric conditions: *NIC_PSYCH→GI_* = 0.11, *NIC_GI→PSYCH_* = 0.06; *NIC_PSYCh→MSK_ =* 0.14, *NIC_MSK→PSYCH_* = 0.13; *NIC_PSYCH→CIRC/HEM_* = 0.17, *NIC_CIRC/HEM→PSYCH_* = 0.05; *NIC_PSYCH→RESP_* = 0.16, *NIC_RESP→PSYCH_* = 0.08; *NIC_PSYCH→ID_* = 0.21, *NIC_ID→PSYCH_* = 0.12, *NIC_PSYCH→NEURO_* = 0.24, *NIC_NEURO→PSYCH_* = 0.16; *NIC_PSYCH→ENDO/METAB_* = 0.13, *NIC_ENDO/METAB→PSYCH_* = 0.01. For comparison’s sake, normalized intra-module co-morbidity for psychiatry was 0.38. The pattern of differences between the predicted and predictive effects of psychiatric diagnosis is unlikely to be due to chance alone (mean of differences *NIC_PSYCH→MED_* - *NIC_MED→PSYCH_* = 0.075, P=0.013 from permutation test with 1000 permutations). Overall, the neurology module showed the strongest evidence of co-morbidity with the psychiatric conditions, followed by the infectious disease module and the musculoskeletal module.

**Figure 3:**
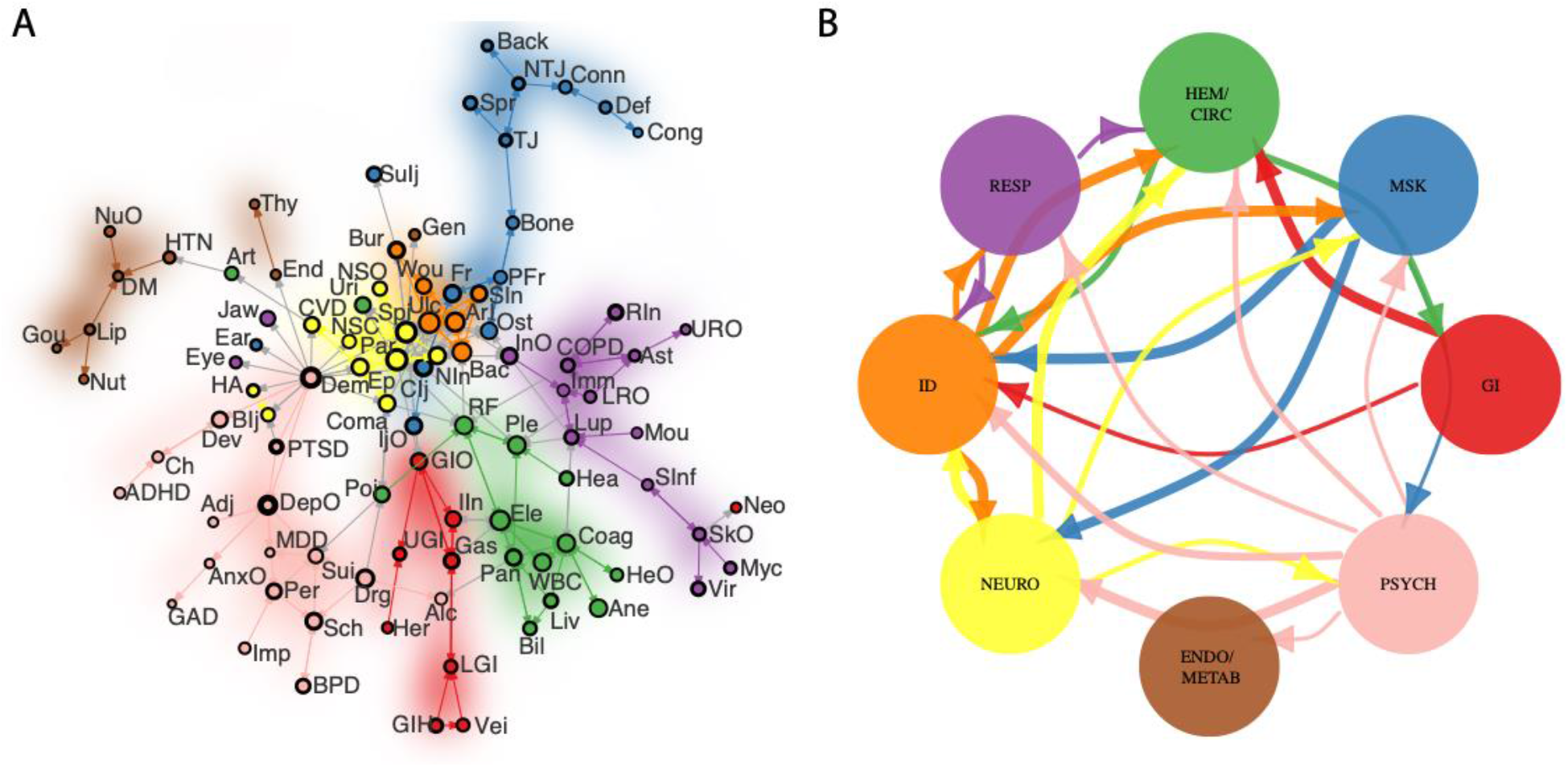
Inter-module and psychiatric-medical co-morbidity. A) Complete network of medical and psychiatric co-morbidity, using the 7-module medical community structure described in Figure 2C and representing the psychiatric conditions as a single module. B) Module-level representation, with edge-weight proportional to the normalized intermodule co-morbidity (NIC, see methods) and showing the top 50% highest-weight edges for the purposes of illustration. For every medical module, *NIC_PSYCH→MED_* > *NIC_MED→PSYCH_*. See Table 2 for list of abbreviations.

The high observed co-morbidity with neurological conditions plausibly stems from a shared basis in brain pathology, but psychiatric conditions’ relatively high co-morbidity with musculoskeletal and ID conditions may be more counterintuitive. Across all psychiatric conditions, substance use disorders have the highest average connectivity with the ID module, suggesting that risk for infectious disease related to intravenous drug use may partially mediate this relationship. On the other hand, MSK co-morbidity may be partially mediated by pain, either due to psychiatric consequences of physical pain or due to a shared neurobiological substrate for physical pain and mental distress. In fact, the association with chronic pain, as measured by the effect size of the chronic pain index on each disorder in the logistic regression model described in the methods, differed between the modules of medically co-morbid conditions =(mean log ratio for MSK module = 0.49, 95% confidence interval (CI) = 0.30-0.73 from 1000 bootstrap samples; NEURO=0.18, CI=0.01-0.38; GI=0.07, CI=0.00-0.18; RESP=0.00, CI=–0.14-0.17; ID = 0.01, CI=–0.04-0.05; ENDO/METAB=–0.02, CI=–0.07-0.04; HEM/CIRC=–0.08, CI=–0.16-0.01; ANOVA predicting pain index by module membership, F=7.6, df=6, p=<0.001). The high association with chronic pain may mediate the co-morbidity between psychiatric and musculoskeletal conditions, and to a lesser extent the co-morbidity between psychiatric and neurological conditions.

### Edge-level associations between medical and psychiatric co-morbidity

Complementary to the above analysis based on community structure, we also investigated specific edges that represented significant co-morbidity between psychiatric and medical conditions. These edges were statistically significant after FDR correction for multiple comparisons (q < 0.05) and also exceeded an odds ratio of 1.5. In the direction of medical conditions predicting psychiatric conditions, 72 edges (5% of possible edges) exceeded the threshold. In contrast, in the other direction of psychiatric conditions predicting medical conditions, 227 edges (17% of possible edges) exceeded the threshold. The following psychiatric conditions were significantly predictive of the largest number of medical disorders: PTSD (15 medical conditions), substance use disorders (26 medical conditions), schizophrenia (29 medical conditions), cognitive disorders (48 medical conditions) and minor depression (71 medical conditions).

The large majority of medical conditions had significant relationships with multiple psychiatric disorders. Underlying the module-level relationship between neurological and psychiatric conditions, particularly strong psychiatric co-morbidity was observed with paralysis, intracranial injury/brain damage, headache, ear conditions, epilepsy/seizures, and cerebrovascular disease. Other broad categories included conditions associated with chronic pain, such as fractures and joint disease, and conditions that could be plausibly explained by behaviors stemming from psychiatric conditions, such as superficial injuries (strongly associated with suicidality/self-harm) and pancreatic conditions (strongly associated with alcohol use). We found statistically significant associations with diabetes, hypertension, and heart disease that have been previously reported and understood either as likely side effects of psychotropic medications^44^, as a function of shared social determinants of health such as homelessness^45^, or due to a shared biological mechanism such as inflammation^46^. The full set of significant co-morbidity relationships is available at https://github.com/aaronab/comorbidity_networks.

### Mortality and Predictive Models

If network-level co-morbidity information is to be useful, it should be associated with clinically meaningful outcomes. A prior study showed a correlation between disease mortality and a measure of co-morbidity strength^47^, and we replicate this finding in our independent dataset (Figure 4). Moreover, we show that this relationship exists for both in-strength and out-strength, although it is stronger for in-strength (Spearman’s *ρ*=0.80, p <0.001, permutation test with 1000 permutations of the relationships between strength and morbidity across conditions) compared to out-strength (Spearman’s *ρ* =0.52, permutation p<0.001; Spearman’s correlation with permutation tests were used out of concern that node strengths were not normally distributed). This relationship also holds true within psychiatric conditions: there was a convergence between conditions with high mortality and those with high in-strength including substance-use disorder, schizophrenia, personality disorders, and suicidality. Minor depression and dementia were outliers in terms of having high out-strength relative to condition mortality. Note that when the psychiatric co-morbidity network was considered separately, there was a small correlation that was not statistically significant between in-strength and out-strength (Spearman’s *ρ* =0.25, permutation p=0.34, Supp. Fig 2A); but when the medical-psychiatry network was considered as a whole, in-strength and out-strength were strongly correlated (Spearman’s *ρ* =0.58, permutation p<0.001, see Supp. Fig 2B). Overall, the extent to which diagnosis with a condition impacts an individual’s risk of death does appear to be related to the condition’s centrality in the comorbidity network.

**Figure 4:**
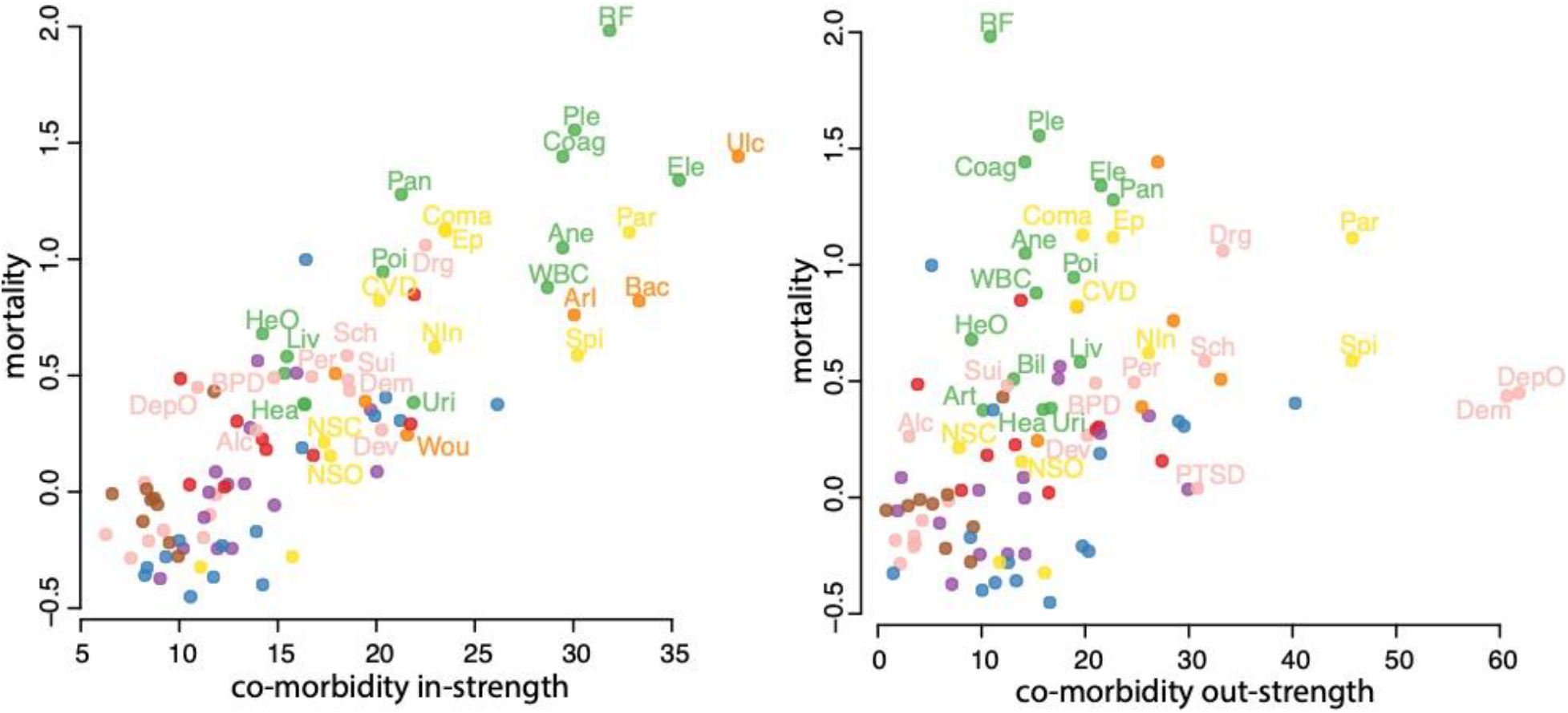
The relation between disease mortality and strength in the co-morbidity network, for in-strength (left) and out-strength (right). Individual nodes are colored according to their community structure per Figure 3. See Table 2 for list of abbreviations.

Age-of-onset of the conditions might be expected to confound the association between mortality and co-morbidity strength, as older people suffer from higher mortality and a greater number of medical co-morbidities. However, age was included in the logistic regression models in order to control for the confound with co-morbidity. Moreover, there was not statistical support for an association between a condition’s age-of-onset (median across all affected individuals) and the condition’s co-morbidity strength (Spearman’s *ρ*=-0.14, permutation p=0.16) nor between age-of-onset and mortality (Spearman’s *ρ*=-0.01, permutation p=0.95), whereas the partial correlation between co-morbidity strength and mortality remained high when controlling for age-of-onset (partial Spearman’s *ρ*=0.80, estimated with R package *ppcor*^48^). We note that the lack of an association between mortality and age-of-onset may be due to the relatively young age of this cohort compared to the entire population served by the VA.

In practice, co-morbidity information could be used to predict the development of subsequent conditions. Here we show the potential to flag clinically high-risk individuals using co-morbidity alone, as an example of this potential utility. Schizophrenia and suicidality were chosen as exemplars given the enormous clinical significance, number of previous studies and worldwide resources invested in the prediction of these conditions^39,40^. The present dataset yielded moderate predictive capacity for both conditions using lasso regression on the co-morbidity data. For schizophrenia, accuracy was 92% in the test set, with a specificity of 92% and a sensitivity of 88%; for suicidality, accuracy was 91% in the test set, with a specificity of 92% and a sensitivity of 90%. In both cases, network-level co-morbidity greatly increased the accuracy of the models compared to predictions based on demographic information alone (age, sex, race, and years-in-VA), as shown by comparing ROC curves with co-morbidity data to ROC curves without the co-morbidity data (Figure 5).

**Figure 5:**
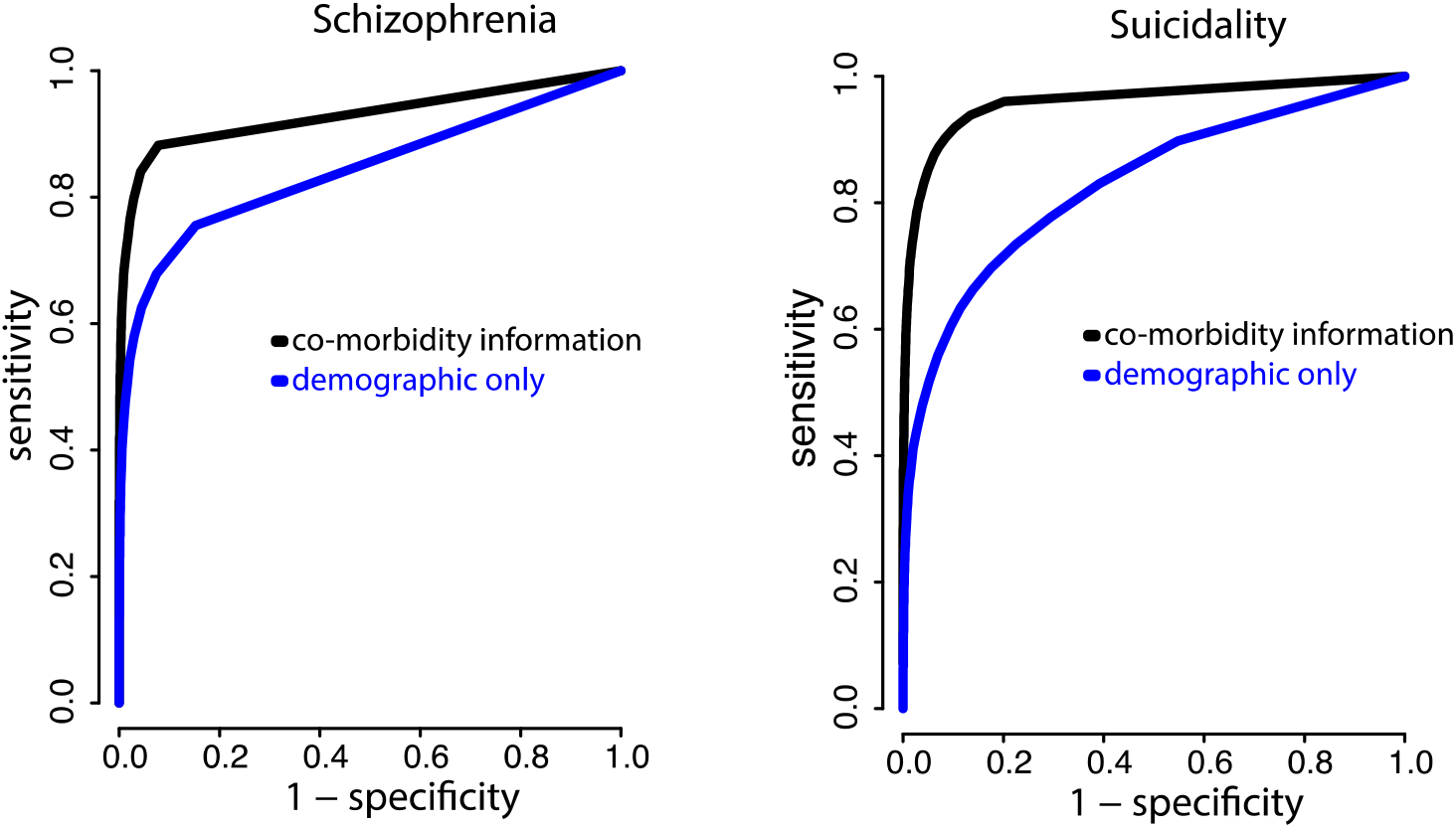
Receiver operating characteristic (ROC) curves for the predication of schizophrenia and suicidality based on the prior (or concurrent) diagnosis with 94 other medical or psychiatric conditions using lasso regression. In blue, ROC curves are shown based on using demographic data only (age, sex, race, and the number of years within the VA health system).

### The impact of biological sex on co-morbidity

Most work to date describes co-morbidity while controlling for the effect of sex, but this process may obscure important sex-related differences in co-morbidity patterns. Here, we show widespread sex-specific augmentation of co-morbidity relationships, with the interaction effect reaching statistical significance in 24% (2,126 out of 8930) of the models fitted across pairs of conditions. As shown in Figure 6A, there was a larger number of condition pairs whose comorbidity was augmented by male sex, as opposed to female sex. Given their high co-morbidity, we more closely considered the sex effects within and between psychiatric and neurologic conditions. The role of PTSD as a predictor node was augmented in male veterans, both with other psychiatric conditions and neurological conditions including headache, brain injury, and epilepsy. In contrast, the role of depressive disorders as predictor nodes was augmented in female veterans (Figure 6B). The full set of significant interaction effects is available at https://github.com/aaronab/comorbidity_networks.

**Figure 6:**
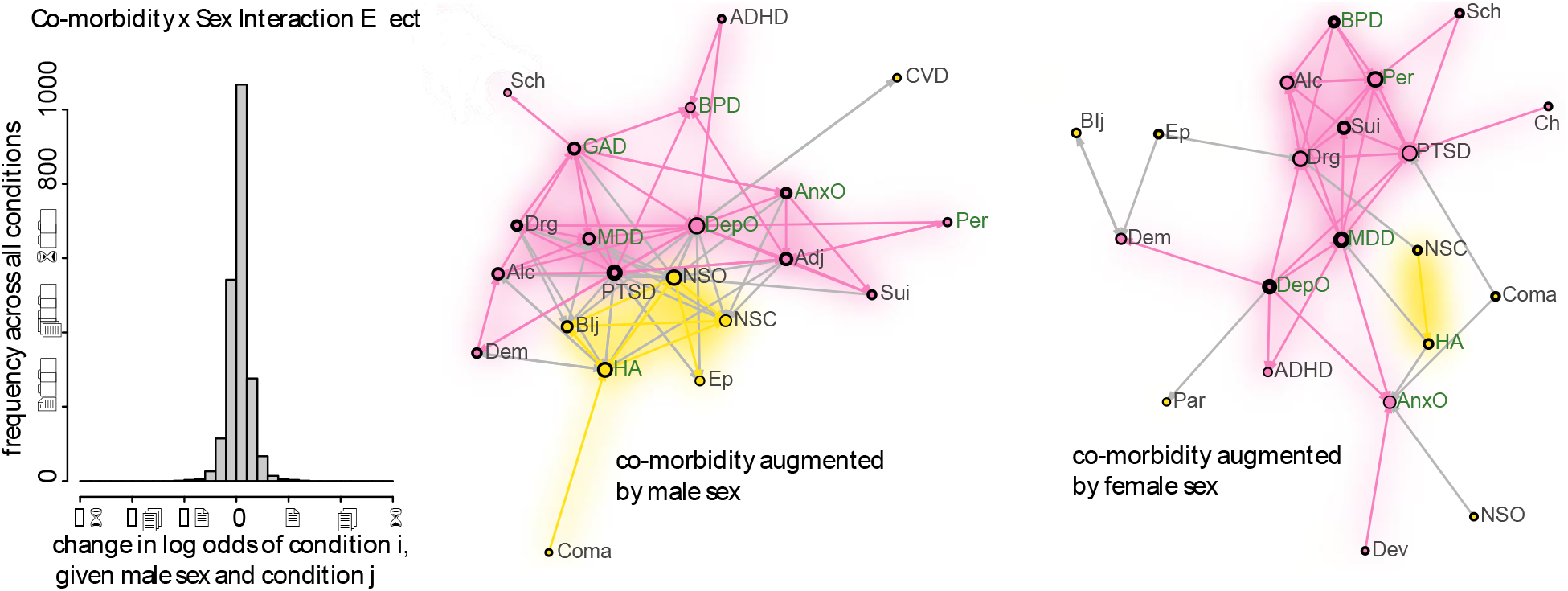
Augmentation of co-morbidity relationships by sex. Histogram of the effect size (log odds increase or decrease) for all statistically significant effects (P<.05 after FDR-correction). Showing a subset of conditions consisting of the psychiatric and neurologic conditions, a plot showing all significant edges whose co-morbidity was augmented by male sex (middle) or by female sex (right). Conditions labelled in green have higher prevalence in female veterans, while conditions labelled in black have higher prevalence in male veterans. See Table 2 for list of abbreviations.

## Discussion

We report the first systematic investigation of network architecture in co-morbid psychiatric and medical conditions in the US Veterans Administration population. We show the multi-scale community structure of co-morbid conditions, with neurological conditions and conditions associated with significant physical pain having a disproportionate co-morbidity with psychiatric conditions. Diagnosis with PTSD or minor depression (depressive symptoms not meeting the threshold for Major Depressive Disorder) strikingly increased the likelihood of future medical morbidity as well as the diagnosis with major psychiatric conditions. In and of itself, network-level co-morbidity information provides a reasonably accurate flag for individuals vulnerable to schizophrenia and suicidality/self-harm. Our results are broadly consistent with previous comprehensive studies of co-morbidity networks^47,49-52^ while extending the past work in multiple ways.

### The biological significance of co-morbidity analysis

Co-morbidity does not inherently stem from a single cause. In any individual, it may be due to chance, although at the population level co-morbidity is vastly higher than predicted by random overlap of independent conditions. This non-independence could be attributable to a variety of causes, including direct causality (alcohol use and pancreatic disease) and shared environmental risk factors (cerebrovascular and heart disease)^53^. Shared genetic risk has been demonstrated for many commonly co-morbid conditions^13-15^, and co-morbidity may also be mediated by underlying psychological vulnerabilities such as impaired distress tolerance^54^. It is plausible that organ systems with greater functional integration across physiological states^55,56^ manifest greater co-morbidity in pathological states. A shared basis in disruption of brain circuitry substrate likely mediates the strong relationship between neurological and psychiatric conditions^57^, conditions in which many similar alterations in the functional and structural properties of brain networks have been reported^58,59^. Meanwhile, the association between pain and psychiatric morbidity has been hypothesized to be related to mutually reinforcing disruption of functional status^60,61^ and to a neurobiological basis of physical and emotional distress^62,63^.

Access to care and diagnostic expertise can certainly influence co-morbidity patterns, such as screening for PTSD at VA clinics potentially increasing co-morbidity with medical conditions. However, PTSD’s association with co-morbid psychiatric and medical illness is not specific to US veterans, having been reported in other international populations^64^. It is unlikely that increased rates of co-morbidity between psychiatric and medical conditions are driven by increased utilization alone, as veterans with psychiatric illness are likely to have fewer medical visits and increased rates of undiagnosed disease^3,65^. Thus, there is reason to believe that medical-psychiatric co-morbidity is likely to be, if anything, under-represented by the current results. Sex-related differences in co-morbidities require further investigation and could result from a mixture of cultural factors, bias among medical practitioners, differential exposure to risk factors, and physiological differences.^4,21^ In general the present methodology cannot cleanly disambiguate patterns of underlying physiologic co-morbidity from patterns in the diagnostic behavior of clinicians, to the extent that these patterns diverge.

Research into the etiology of co-morbid conditions is hampered by the intentional exclusion of individuals with more than one diagnosis from basic scientific investigations^66^. However, individuals with specific co-morbidity patterns may actually be enriched for specific risk factors of biological interest. Neuroinflammatory processes may mediate co-morbidity between PTSD, MDD, and substance use disorder^67^, as well as the link between these disorders, cardiovascular risk^68^ and pain sensitivity^69^. Overlapping alterations in glutamate and GABA systems may affect PTSD, MDD, and substance use disorders^70^. Rather than being excluded, it may be that individuals with multiple co-morbidities should become a focus of future research.

### Co-morbidity, clinical practice and public health

Information from network models of population co-morbidity has the potential to improve diagnostic accuracy and impact clinical decisions^18^. Evidence-based diagnosis is often described in terms of the pre-test probability for a condition, which is updated based on the results of specific diagnostic tests^71^. In this framework, information about existing medical and psychiatric conditions could markedly increase diagnostic accuracy, and this information is readily available from EHRs. Our results demonstrate the potential utility of such a procedure in the VA population.

Given the tendency to exclude patients with co-morbidities from research, it is perhaps not surprising that co-morbidity has a disproportionate public health impact. Indeed, medical ‘multimorbidity’ is increasingly understood as a public health crisis^1^, and this burden is likely increased when considering the relationships between medical and psychiatric multimorbidity^3^. In fact, a prior network analysis of Medicare data showed that diseases with more co-morbid connections (greater co-morbidity with other diseases than expected by chance alone) also tend to have higher rates of mortality^47^. An understanding of the complex relationships between psychiatric conditions, and their interactions with medical illness, could lead not only to earlier and more accurate diagnoses of co-morbid conditions but to the development of targeted treatment interventions.

As a specific example informed by the current study, the large number of veterans with co-morbid PTSD, substance use disorders, chronic pain, and/or neurological disease are a potential target for more individualized treatment approaches. The co-morbidity between these conditions is remarkably high, and proper identification of these patients may facilitate investigations of medications with dual benefit, e.g. serotonin-norepinephrine reuptake inhibitors to target both PTSD and pain, or buprenorphine to target both pain and opioid use disorder. It is important to note that the common-treatment approach often lacks empirical support, and efficacy may be decreased in the presence of co-morbidity. Individuals with medical and psychiatric multimorbidity may represent a subgroup requiring alternative treatment strategies^4^. There are encouraging trends in this direction, including the investigation of psychotherapeutic strategies for co-morbid PTSD and substance use disorder^72^, as well as pharmacological interventions^73^ and interventions at the level of healthcare systems^74^.

### Methodological considerations and future directions

Several methodological considerations related to the present analysis are worthy of note. Estimated frequency of multimorbidity varies substantially across studies, depending on factors such as the nature of the population (for example, primary care or population-based surveys) and the number of clinical conditions assessed. The present findings may not extend to all US veterans who served in Iraq and Afghanistan, as the data are derived only from people who actually received care in the VA system. In addition to the large political, cultural, and economic significance of healthcare outcomes of veterans, VA healthcare provides a lens into complex health systems in general. However, unique characteristics such as rates of psychological trauma may limit external validity.

Mapping between ICD codes and clinical conditions is a significant limitation, as the accuracy of these mappings is only approximate^75^. Although our approach to mapping has been validated in other datasets^21-23,36^, the possibility remains that diagnostic bias - as opposed to underlying pathophysiology - also influences co-morbidity patterns. We chose to focus on an intermediate level of specificity in terms of medical conditions, level 2 within the four levels of AHRQ mappings, which limits our findings to this intermediate level. One possible area for future research would be to use a multilayer network approach^76^ to map co-morbidity relationships across levels simultaneously, which would allow us to test the generalizability of findings to different diagnostic resolutions.

The present approach investigates only first-order co-morbidity. It does not speak to whether diagnosis with specific prior conditions can influence co-morbidity between two other conditions. This could be addressed in future work by iteratively constructing co-morbidity networks for the subset of individuals with different conditions (for example, PTSD versus PTSD and depression). Another related possibility is that the underlying structure could be more appropriately modelled as a hypernetwork, wherein hyperedges connecting more than just two conditions are the basis for the network.^77^

Future work has the potential to greatly extend and clarify this study. Conditions such as homelessness, employment status, and social isolation, which are known to interact with morbidity of mental and medical illness, could be incorporated into future analyses. In addition, technical developments promise greater precision in terms of defining co-morbid relationships, investigating temporal relationships, and isolating causality from correlation^78^. An important area of investigation is the convergence and divergence of different models of the ‘diseasome’^79,80^; for example, comparing networks derived from clinical co-morbidity, symptomatology genetic risk, social vulnerability, pharmacological response profiles and (in the case of mental illness) neurobiological substrates.

### Conclusions

The present study provides support for a network perspective on psychiatric and medical comorbidity. Systematic relationships, such as the marked predictive power of psychiatric conditions in terms of future medical morbidity, yield insights into how multimorbidity unfolds over time and how individuals interact with healthcare systems. Commonly co-morbid conditions, identified by data driven approaches, represent novel phenotypes for both scientific investigation and also clinical intervention.

## Data Availability

Data associated with this article is available at is available on GitHub.

https://github.com/aaronab/comorbidity_networks.

## Acknowledgements

We are grateful to Bob Rosenheck and Joe Erdos for helpful feedback on earlier drafts of the manuscript. We are grateful to Rick Betzel for code to visualize community-labeled networks. The views expressed in this article are those of the authors and do not necessarily reflect the position or policy of the U.S. Department of Veterans Affairs.

